# Emergency department presentations of community-acquired lower respiratory tract disease in Bristol, UK: a prospective cohort study 2022-2023

**DOI:** 10.1101/2025.02.10.25321388

**Authors:** Catherine Hyams, Robert Challen, Maria Lahuerta, Elizabeth Begier, Serena McGuinness, Madeleine Clout, Jo Southern, James Campling, Jennifer Oliver, Christian Theilacker, Gillian Ellsbury, Nick Maskell, Bradford Gessner, Leon Danon, Adam Finn, the AvonCAP Research Group

## Abstract

**Background:** Recent reports highlight the importance of acute lower respiratory tract disease (aLRTD) for patients, but data describing incidence and burden in emergency departments (ED) are lacking.

**Methods:** A cohort study ascertaining cases prospectively at two EDs in Bristol, UK, enrolling adults (≥18years [y]) presenting with aLRTD from 01-Aug-22 to 31-Jul-23. Multivariate logistic regression modelled risk of hospitalisation. Incidence was estimated per 1000 person-years, using adult population estimates for AvonCAP study catchment area.

**Results:** 151865 ED visits, with 9452 (6.2%) aLRTD cases: 2376 (25%) were discharged and 7076 (75%) subsequently hospitalised, including: 3663 (38.8%) pneumonia, 4167 (44.1%) non-pneumonic lower respiratory tract infection (NP-LRTI) and 1622 (17.2%) cases without evidence of infection. Univariate analysis demonstrated aLRTD patients discharged were younger than those hospitalised (median age 43.4y, IQR:29.4-62.3 versus 74.0y, IQR:59.8-83.5), and less likely to have pneumonia (17.0% versus 46.0%, respectively). Smoking, heart failure at presentation and underlying chronic cardiac disease conferred risk of admission, above an age effect in the adjusted logistic regression model.

Total ED aLRTD incidence was 12.8/1000 person-years (9.6 admitted, 3.2 seen and discharged), with incidences of 7.0 and 36.8/1000 person-years in 18-64y and ≥65y, respectively, and incidence increased with patient age: 39.5 and 82.5/1000 person-years in 75-84y and ≥85y age groups, respectively.

**Interpretation:** We report a higher ED aLRTD incidence than in recent British Thoracic Society and Getting It Right First Time reports. This is concerning, particularly in older adults, and may be reduced by respiratory disease optimisation and public health initiatives including smoking cessation and vaccination programmes.

**KEY MESSAGES:** *What is already known on this topic:* The incidence of acute lower respiratory tract disease (aLRTD) is high in secondary care settings, particularly in emergency departments (ED), with the BTS and Getting It Right First Time reports showing increasing incidence before the emergence of SARS-CoV-2.

*What this study adds:* This study provides accurate population-based estimates of the incidence of aLRTD seen in ED following the emergence of SARS-CoV-2, as well as detailed descriptions of these patients, stratified by requirement for hospital admission.

*How this study might affect research, practice or policy:* These data and analyses are essential to inform public health decision-making and planning to ensure the appropriate resource allocation and treatment for patients with aLRTD. They highlight the importance of chronic underlying cardiac and respiratory disease as risk factors for aLRTD and admission, and patient outcomes may be improved through disease optimization, as well as smoking cessation and vaccination programmes.

## INTRODUCTION

Acute lower respiratory tract disease (aLRTD) encompasses pneumonia and non-pneumonic lower respiratory tract infection (NP-LRTI), chronic respiratory disease exacerbation (CRDE) and heart failure (HF) [1]. aLRTD remains a significant cause of morbidity and mortality in adults [2], despite multiple established vaccination programmes and other public health measures to reduce disease. Only considering primary diagnoses, respiratory disease is the second most commonly recorded presenting complaint in Emergency Departments (ED), following dislocation/fracture/joint injury/amputation [3]. ED attendances for respiratory diseases have increased in the UK, accounting for 2.5% (463,511 cases) and 3.8% (913,646 cases) of ED attendances in 2010-11 and 2017-18, respectively, with a 31% increase between 2015-16 and 2016-17 [3], [4]. The Getting it Right First Time (GIRFT) analysis of Emergency Care Data System (ECDS) data reported similar findings in 2019/20, finding that respiratory diseases were the most common cause of ED attendance and that respiratory infection was the third most common ED discharge code [5]. The rate of ED attendance for respiratory disease has risen at three times the rate of all visits over the past seven years [2]. In contrast, the Nuffield Trust found that ED attendances for chronic obstructive pulmonary disease (COPD) and asthma did not vary significantly between 2006 and 2019, but reported fewer emergency admissions pertaining to these conditions in 2020, aligning with the start of the COVID-19 pandemic [6,7]. Notably, the seasonal variability in respiratory presentations to ED is far greater than in many other disease areas; in the UK in 2016, respiratory hospital admissions resulting from ED attendances almost doubled from a low point in August to a peak in December [7].

High quality clinical data collection is imperative to ensure the quality and accuracy of analytical results; however, applying rigorous diagnostic criteria to achieve these results in clinical practice is often not feasible, even for common conditions. Countries such as the UK [8], France [9], and US [10] have adopted syndromic surveillance using routine electronic emergency care data to monitor and rapidly detect changes in healthcare resource usage [11]. Since 2017, ED activity has been reported in the UK via the national data set for urgent and emergency care (ECDS) [12]. The British Thoracic Society analysed ECDS data in 2018 [3], GIRFT in 2021 [5] and The Nuffield Trust used Hospital Episode Statistics (HES) data [6]. Other sources of UK respiratory infection surveillance data include laboratory surveillance for specific pathogens, including community surveillance of severe acute respiratory syndrome coronavirus 2 (SARS-CoV-2), respiratory syncytial virus (RSV) and influenza using surveillance methods such as HPZone; primary care surveillance methods, such as the Royal College of General Practitioners (RCGP) Weekly Returns Service; RCGP influenza-like-illness sentinel swabbing and serology; and, secondary care surveillance methods, including UKHSA Second Generation Surveillance System (SGSS), SARI-watch and Severe Respiratory Failure centres [13]. While these data and reports all highlight the impact of lung disease on ED attendance and hospital admissions, they lack granularity around the cohort of patients with respiratory disease seen in ED, and do not describe differences between admitted vs discharged patients.

Because the impact of aLRTD on hospital services is significant, particularly during the winter months, and appears to be increasing [5] with little or no improvement in outcomes for people with lung disease in the UK for more than ten years [14], we undertook a prospective cohort study to determine the incidence of aLRTD in EDs at two hospitals in Bristol, UK. We used active case ascertainment methods, considered the “gold standard” in public health surveillance [15], to describe individuals presenting for emergency care. These data were collected using the same methodology as ongoing work in primary (ISRCTN:64997989) and secondary care (ISRCTN: 17354061), which will allow these analyses to be combined to provide an estimate of aLRTD incidence across an entire healthcare network.

## METHODS

### Ethics and permission

The study was approved by the Health Research Authority Research Ethics Committee East of England, Essex, reference 20/EE/0157, ISRCTN:17354061.

Informed consent was obtained from cognisant patients, and declarations for participation from consultees for individuals lacking capacity. If it was not practical to approach individuals for consent, data were included using approval from the Clinical Advisory Group under Section 251 of the 2006 NHS Act.

### Patient and Public Involvement

Patients or the public were not involved in the design, or conduct, or reporting, or dissemination plans of our research.

### Study design

This is a prospective observational cohort study of adults seen in the ED of two large university hospitals in Bristol, UK. All adults (aged ≥18 years, [y]) attending the ED at both participating hospitals between 1-Aug-2022 and 31-Jul-2023, were screened for aLRTD as previously published [1]. Patients were screened for signs/symptoms of respiratory disease and those with either a clinical/radiological diagnosis of aLRTD, or ≥2 signs/symptoms consistent with aLRTD and disease ≤28 days in duration, were included. Signs and symptoms included documented fever (≥38°C) or hypothermia (<35·5°C); cough; increased sputum volume or discolouration; pleurisy; dyspnoea; tachypnoea; examination findings (e.g. crepitations); or radiological changes suggestive of aLRTD, such as consolidation or pulmonary oedema. Patients were excluded if their symptoms developed ≤7 days from discharge from a hospital admission to exclude hospital-acquired infection. Patients with aLRTD signs/symptoms but a confirmed alternative diagnosis were excluded (e.g. fever and tachypnoea attributable to urosepsis). Eligible cases of aLRTD disease were classified to the different aLRTD subgroups (pneumonia, NP-LRTI, and non-infective aLRTD i.e. HF and CRDE with no infective component) following the case definitions provided below and in Supplementary Data 1.

As an observational study, all patient management and investigation was undertaken at the discretion of the treating physician. Demographic and clinical data were collected from patient medical records in a standardised electronic clinical record form (eCRF) using REDCap [16]. We collected data on co-morbidities at ED attendance and obtained vaccination records from linked general practitioner (GP) records. We also determined the Charlson comorbidity index (CCI; with published estimates of 10-year survival) [17] and CURB-65 score [18] for each case.

### Case definitions

Full case definitions can be found in Supplementary Data 1. Acute lower respiratory tract disease (aLRTD) encompasses pneumonia, lower respiratory tract infection (LRTI), acute bronchitis, exacerbation of underlying respiratory disease including asthma and chronic obstructive pulmonary disease (COPD). Pneumothorax, pulmonary embolism, progression or new diagnosis of primary or secondary lung malignancy were excluded from aLRTD. Pneumonia was classified as acute respiratory illness with confirmed radiological changes compatible with infection or if the treating clinician confirmed the diagnosis. In keeping with National Institute for Health and Care Excellence (NICE) and British Thoracic Society (BTS) guidelines [19,20] patients assigned a pneumonia diagnosis were counted as a pneumonia case even in the absence of radiological investigation or infiltrate on imaging, due to known false-negative radiology in pneumonia. NP-LRTI was defined as the presence of signs and symptoms of acute lower respiratory tract infection in the absence of radiological change consistent with pneumonia or a clinical diagnosis of pneumonia. Under these case definitions, any patients with aLRTD signs or symptoms due to non-infectious aLRTD (i.e. no evidence LRTI) would have been assigned to CRDE or HF groups but not NP-LRTI or pneumonia. Thus no evidence of LRTI included non-infective exacerbations of chronic respiratory disease (e.g. asthma, COPD) and HF where there is no associated pulmonary infection component.

### Outcomes

We determined whether each case was admitted to the care of an inpatient specialty team (i.e. admitted to hospital) and proportions for subsequent presentation (all-cause) to ED at 7 days and 30 days following initial attendance. The 2019 base AvonCAP study population for incidence rate calculations was estimated, as previously described, including full methodology [21]. Briefly, hospital admission data were linked to aggregated GP patient registration data within NHS Bristol, North Somerset and South Gloucestershire Clinical Commissioning Group for 2017–2019. The proportion of GP practices’ aLRTD hospitalisations that occurred at a specific study hospital was multiplied by their patient registration count for six age groups to obtain the practices’ contribution to that hospital’s denominator. The age stratified 2019 base population was adjusted for changes in the demographic profile of GP registrations due to migration and the ageing population, to give time varying estimates of the base population, as described in detail in Supplementary Methods.

### Statistical Analysis

The primary goal of this analysis was to report incidence rates by clinical presentation of aLRTD in the ED to inform public health priorities against a background of evolving epidemiology. Secondary objectives included comparing the patients admitted to inpatient care with those discharged from ED, and describing any difference in severity between these two patient groups.

Characteristics of all aLRTD ED visits are described with categorical data presented as counts and percentages, and continuous data as appropriate either as means with standard deviations (SD) or medians with interquartile (IQR) ranges, with normality of distributions determined with the Anderson-Darling test. aLRTD was stratified into cases that present clinically with 1) pneumonia, 2) NP-LRTI or 3) with no evidence of lower respiratory tract infection. These three categories are disjoint, but may co-present with either heart failure or exacerbation of chronic respiratory disease, or both. Pneumonic and NP-LRTI groups may have had evidence of SARS-CoV-2 as a causative pathogen, determined by PCR (polymerase chain reaction) or lateral flow device.

All persons aged ≥18y contributed to the denominator for incidence estimate calculations which are reported per 1000 person years for the whole cohort (incidence per 100,000 people per year is presented in Supplementary Results), and stratified by clinical presentation and admission status. Time varying and age stratified denominator sizes were estimated as described in the Supplementary Methods. Time varying estimates of admission incidence from weekly case counts were made using maximum likelihood, assuming the observed case count is a binomially distributed quantity with a time varying probability, with the time varying denominator as the sample size. The weekly probability was estimated using a locally fitted order 2 polynomial using a logistic link function using the methods of Loader *et al*. [22], and converted to an incidence per 1000 patient years by multiplication. A sensitivity analysis was conducted using different base population models as described in the Supplementary Methods. Crude case-fatality risk within 2 days, within 30 days and risk of direct hospital admission from ED, are presented, stratified by clinical presentation, for the whole data set; time spent in the ED, and risk of delayed admission within the 30 days following ED discharge are calculated for the subgroup of patients not admitted; hospital length of stay and, intensive care unit (ICU) admission rates for the subgroup of patients admitted. A logistic regression model was fitted using maximum likelihood estimation to assess the factors associated with the risk of hospital admission in patients seen in ED. Variables potentially related to admission were selected a-priori and univariate regression performed. Age was observed as the major determinant and a second set of regressions for every a-priori covariate minimally adjusted for age were performed to identify major interactions. A fully adjusted model containing all covariates was then used to identify variables with residual significant effect on hospital admission. A final parsimonious adjusted model was constructed with only those variables with remaining significance. All analyses were conducted using R [23].

## RESULTS

Of the 151,865 adult patients presenting to the two Bristol EDs between 1st August 2022 and 31st July 2023, 9,769 (6.4%) met the case definition of aLRTD. After removing readmissions for the same illness episode (ie, attendances within 7 days of each other) and those that declined consent, there were 9,452 (6.2%) distinct episodes of aLRTD. Of these, 2,376 (25%) were discharged from ED and 7,076 (75%) were admitted under an inpatient specialty team (Figure 1). Overall, 52.1% of patients were female, with a median age 69.2y IQR [48.6—81.1] and 64% had a CCI score ≥3. There were 3,663 cases of pneumonia (38.8% of all aLRTD ED visits), 4,167 NP-LRTI (44.1% aLRTD visits) and 1,622 cases with no evidence of LRTI (17.2% aLRTD visits). There were 7,575 (80.1%) cases of mild disease (CURB-65 0-1), 1,680 (17.8%) moderate (CURB-65=2) and 197 (2.1%) cases of severe disease (CURB-65 ≥3) (Table 1).

**Figure 1:**
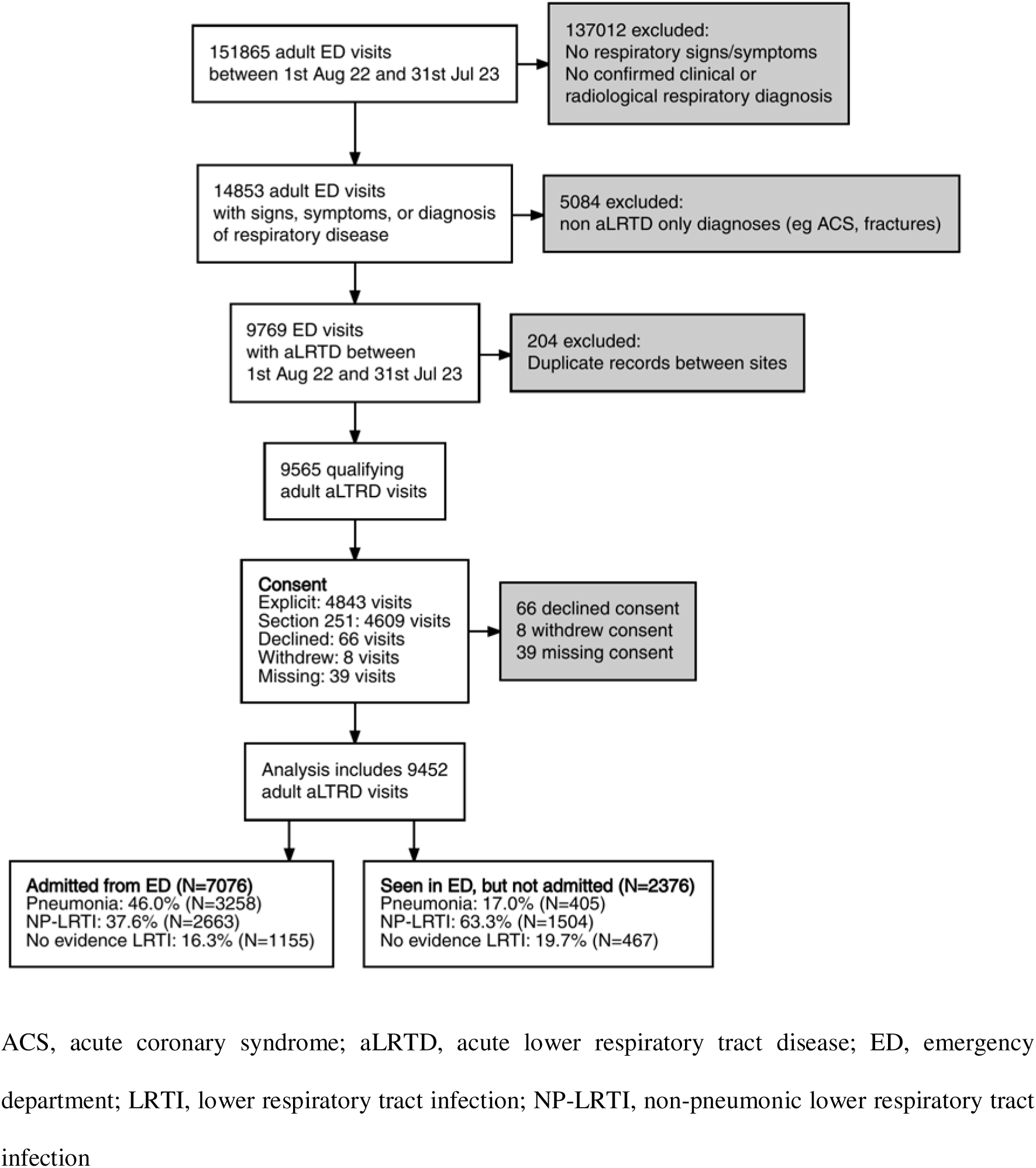
Study Flow Diagram.

On univariate analysis, patients who were seen in ED, but not admitted to hospital, were younger (median age 43.4y, IQR 29.4-62.3 versus 74.0y, IQR 59.8-83.5); were less likely to reside in a long-term care facility I(2.4% versus 8.5%); less likely to have pneumonia (17.0% versus 46.0%); and had less comorbid disease (CCI ≥3 of 27.6% versus 76.3%), in comparison to those who were admitted (Supplementary Data 2). As expected, patients with moderate or severe disease were more likely to be admitted (Supplementary Data 2), and those with pneumonia were more likely to be hospitalised than those with NP-LTRI or no evidence of LRTI (88.9% versus 63.9% and 71.2%, respectively) (Table 2). Patients seen in ED with aLRTD had a mortality of 1.5% within 2 days of presentation and 6.9% within 30 days of presentation, and those discharged from ED had a mortality of 0.5% and 1.1% within 2 and 30 days of presentation, respectively (Table 2). In keeping with the increased hospital admission rate, patients with pneumonia also had a longer hospital admission, were more likely to require intensive care unit (ICU) level care and, even if they were not admitted, had a longer attendance in ED compared to patients with NP-LRTI and those with no evidence of LRTI (Table 2).

The risk of hospital admission resulting from an ED visit was heavily dependent on age. In an unadjusted comparison, adults ≥85y were 26 times more likely to be admitted than those aged 18-34y (Supplementary Data 4); other variables which were closely correlated with age contributed to this increased risk. In an adjusted logistic regression model (Table 3) we found that smoking, and specific pre-existing comorbidities, underlying chronic heart disease, type 1 diabetes, and presentation with acute HF inferred significant risk, over and above the effect of age. Patients with NP-LRTI or non-infective causes of aLTRD had 60% lower odds of being admitted to hospital (OR=0.41; 95% CI 0.35–0.47 for NP-LRTI) compared to those with pneumonia. Patients with cough were less likely to be admitted; patients with confusion were at high risk of admission, as were those who had any oxygen requirement in the ED.

The incidence of aLRTD in EDs in Bristol is shown in Figure 2. Incidence increased with patient age, with particularly high incidence in those aged 75-84y and ≥85y. The time series showed a similar pattern across all age groups, with cases increasing from August 2022 and plateauing in November 2022 until a surge in cases in December 2022, peaking at an instantaneous incidence rate of 147 [95% CI 128-169] per 1000 person years (17 Dec) in the ≥85y age group. This was followed by declining aLRTD ED attendances from January to July 2023. The average incidence of ED episodes for aLRTD over the whole period was calculated as 12.8 per 1000 person years across all age groups, with an incidence of 7.0 and 36.8 per 1000 person years in adults aged 18-64y and ≥65y, respectively (Table 4). Incidence estimates were affected by assumptions on population size and values under different assumptions are shown in Supplementary Data 5, and the results of viral testing on patients with aLRTD presented in Supplementary Data 6

## DISCUSSION

This prospective cohort study reports the incidence of aLRTD in ED in Bristol, UK, over a 12-month period occurring after a period during which non-pharmaceutical public health interventions were nationally implemented to control the circulation of SARS-CoV-2, and these may have changed respiratory infection epidemiology. It is thus important that acute respiratory disease is described following the emergence of SARS-CoV-2 and the resultant pandemic, allowing accurate ascertainment of burden and incidence, enabling appropriate public health planning to be undertaken. We found community-acquired aLRTD in adults represented 6.2% of adult attendances in EDs in Bristol: 82.9% (n=7830) of these cases were associated with respiratory infection (5.2% all ED cases). Notably, 74.9% of aLRTD patients attending ED required subsequent hospital admission and we found an all-cause mortality of 1.5% and 6.9% within 2 and 30 days of presentation to ED with aLRTD, respectively, and among those discharged 1.1% died within 30 days. Ascertaining the total secondary care burden of aLRTD requires combining the patients seen and discharged in ED, as well as those admitted from ED and directly to an inpatient specialty team. A previous analysis reports data on aLRTD requiring inpatient care from the same hospitals as this analysis, albeit over a different time-period encompassing the COVID-19 pandemic [1]. We find that the additional burden of aLRTD seen in ED but not admitted may increase the estimated total aLRTD in secondary care by 25% more than previously reported, highlighting the impact of aLRTD on healthcare systems [1]. Additionally, the total burden of aLRTD will include cases seen in primary care and those not requiring medical treatment, which will lead to further direct and indirect costs, including lost working days and decreased quality of life. Overall, the incidence estimates we provide highlight the continued burden of aLRTD in EDs following the COVID-19 pandemic, at which time it was reported that ED attendances due to some respiratory conditions had decreased [6]. To our knowledge, these are the first population-based incidence estimates of aLRTD and its subgroups in EDs.

We highlight the very high burden of acute respiratory disease in ED, with the 6.2% observed in this cohort being higher than previously reported estimates, and incidence estimates for overall disease being 2.8, 4.0 and 5.6 per 1000 person yrs in adults aged 65-74y, 75-84y and ≥85y, respectively. In context, GIRFT reported for 2019/20 2.5% of ED attendances were due to LRTI, 2.4% upper respiratory tract infection, 0.92% asthma, 0.91% COPD, 0.56% bronchiolitis and 0.45% bronchopneumonia among all patients, including children [5]. In 2020, national data reported 13.96% of UK ED attendances were due to cardio-pulmonary conditions [24]: higher than our estimate but including cardiac cases, which we excluded. The US National Hospital Ambulatory Medical Care Survey found 10.6% of ED visits in 2017 were attributable to respiratory disease [25]. Our results show similar temporal trends to those in UK National ED syndromic surveillance system [26] and BTS review of the impact of lung disease [3], highlighting the pressure on healthcare systems, particularly EDs, due to respiratory illness during the winter months. Increased community transmission of respiratory pathogens such as influenza, RSV, SARS-CoV-2, pneumococcus would lead to more acute episodes of infection, and consequently more patients needing care across all levels of healthcare. National surveillance data from the UKHSA showed a rise in circulation and cases of respiratory infection in November, that peaked in January, and declined by March [27] aligning with the results presented here; we found a rise in SARS-CoV-2 throughout winter, with a rise in influenza A and B in December and February-March, in line with national surveillance (Supplementary Data 6). Our study provides data describing the patients requiring treatment in ED for aLRTD and further stratifies the cohort into patients by age and whether they required subsequent hospital admission. This enables us to provide population-based summary incidence estimates which are essential to inform public health decision making and planning. Additionally, our analysis identifying smoking, specific pre-existing comorbidities including underlying chronic heart disease and type 1 diabetes over and above the effect of age may enable identification of strategies to improve the health of these at-risk patients and not only improve their outcomes but mitigate their risk of aLRTD.

Overall, 48.1% of this cohort experienced an exacerbation of an underlying chronic respiratory condition; on univariate analysis, we found patients with asthma were more likely to be discharged from ED than admitted; conversely, COPD patients were more likely to be hospitalised than discharged from ED (Supplementary Data 2). The impact of underlying respiratory disease in the UK is highlighted by data reporting an age-sex standardized hospital admission rate that is either higher than average or the highest found when compared to comparator countries [6,28]. Despite the significant economic burden on the National Healthcare Service (NHS), estimated at £1.9 billion and £3 billion for COPD and asthma, respectively [2], hospital admission rates have not varied and lung disease including infection remain manifestations of inequality [29,30]. The Taskforce of Lung Health’s five-year plan [14] highlighted improving outcomes for patients with COPD and asthma as a public health priority: this analysis confirms these patients still have a high disease burden following SARS-CoV-2 emergence and these patients remain at risk of poor outcomes. We found that smoking conferred a risk of admission over and above the effect of age, highlighting the importance of smoking cessation programmes as well as other public health initiatives and evidence-based interventions. Regarding patient outcomes over time, we noted that the mortality rates we observed in pneumonia are higher in this study than those estimated in the original CURB-65 mortality risk, which estimated 30-day mortality of 1.5%, 6.8% and 9.6% in patients with a CURB-65 score 0-1, 2 and overall, respectively. We found the mortality risk of pneumonia at 30-days was 14.1% in this patient cohort, and this may reflect the increasing age, burden of pre-existing comorbid disease and frailty in this patient population in comparison to the cohort in which the CURB-65 mortality risk was assessed [18].

We provide a highly detailed description of aLRTD patients attending EDs, stratified by requirement for hospital admission, while recent reports from BTS [3], GIRFT [5] and Nuffield Trust [6] all lacked granularity in their descriptions of this patient group. These data are important in underpinning and informing change in the care and treatment of patients with respiratory disease. Older patients with pneumonia, and those with heart failure were at high risk of hospital admission. Overall, 34.5% of those admitted did not require supplementary oxygen and 23.8% had a CURB-65 score ≤1, although 44.1% were classified as frail and 52.4% had a CCI score ≥3 (Supplementary Data 2). Thus, underlying patient frailty or comorbidity, which increases with age, probably contributes to the admission requirement of patients with aLRTD. We found that patients with cough were less likely to be admitted; possibly inability to cough being a proxy for frailty, which would be aligned with our previous findings that elderly patients with aLRTD presented atypically and were less likely to cough [31]. We did not find evidence within this cohort that, once in ED, socioeconomic deprivation affected the risk of hospitalisation (Supplementary Data 4). When combined with other reports indicating that patients were more likely to attend ED with respiratory conditions [30,32], and that socio-economic deprivation was associated with worse respiratory diseases [33–35], this suggests that poorly controlled disease may be driving ED attendance of these patients. Importantly, management of long-term conditions is undertaken and coordinated by primary care in the UK, thus patients circumventing primary care by visiting EDs may not benefit from optimisation of underlying chronic respiratory conditions, leading to continued deterioration and repeated ED attendance.

This study has many strengths. Firstly, it was undertaken prospectively and by screening ED admissions for aLRTD signs/symptoms. Prospective, comprehensive case ascertainment within a defined geographical area remains the gold standard epidemiological methodology for estimating disease incidence. This study did not rely on coding or solely on national data-linkage, assessing each case individually and gathering complete data. Importantly, the NHS provides healthcare free at the point of access; therefore, ED attendance is not associated with direct cost to the patient, which may bias patient populations where private healthcare or insurance-based medical systems apply. We had ethical approval to enroll all adults seen in EDs: our cohort was therefore comprehensive and not subject to enrolment biases, ensuring patients lacking capacity, such as those severely ill or with advanced dementia or other frailty, were not under-represented in this study. This study was conducted at two hospitals which provide all acute ED care for the same city and time-period to undertake comprehensive surveillance in a defined geographical area with a well-defined local population, and therefore provide an accurate estimate of disease incidence and severity. The medical records were linked with community records to obtain detailed and accurate data for each study participant. Finally, we calculated incidence using a denominator derived from GP records and hospital utilisation data, providing increased accuracy compared to population estimates based on assumptions using local geographic boundaries and their corresponding census data [21].

There are some limitations to this study. Firstly, we conducted case ascertainment over a 12-month period, so cannot report incidence estimates or disease trends over a longer time period. This study occurred after the COVID-19 vaccination programme commenced in the UK, and after periods of social distancing and mandated mask wearing, but we note that some individuals may have continued to wear masks and maintain social distancing, but we do not have data to assess this. Whilst we assessed aLRTD at both acute care NHS hospitals in Bristol, we cannot be sure this is generalisable to other cities and regions. While the population of Bristol is representative of the UK, it is predominantly White-British (84%) [36] and therefore aLRTD disease in cohorts with different ethnicity may vary from that reported here. It is difficult to determine whether access to healthcare changed before or during the study period; for example, clinicians or patients may have preferred treatment at home, thereby affecting severity assessment, ED attendance and hospital admission rates, and our observed incidence. Neither hospital in this study had an attached unit staffed by general practitioners, but both had a same day emergency care unit (SDEC) staffed by inpatient specialists to avoid admissions to either the inpatient service or ED, and this may affect our incidence calculations compared to other hospitals. Patients in this cohort were investigated and treated at the discretion of treating clinicians, and while 72.3% underwent radiology investigation, only 23.2% and 57.1% underwent any form of microbiological or virological testing, respectively, so we cannot describe the pathogens responsible for illnesses resulting in ED attendance in the wider patient cohort, and data on testing for patients not eligible for this study was not available to us. Directly or indirectly, age is a key determinant in the decision to admit patients from the ED [19]. Other factors that are associated with hospital admission are often also determined by (e.g. vaccination) or correlated (e.g. pre-existing comorbidities) with age [1–7]. These factors were excluded from the model as logistic regression would only be able to partly adjust for these interactions. Dissecting the relative effect of age and these linked factors would require further analysis using a matched cohort.

In conclusion, we found individuals who attended EDs with community-acquired aLRTD were elderly, had high frailty and underlying comorbid disease rates, including chronic respiratory disease. We estimated aLRTD incidence to be 7.3 and 38.1 per 1000 person-years in adults aged 18-64y and ≥65y, respectively: finding a higher proportion of ED attendances due to aLRTD than reported by BTS and GIRFT, highlighting the increasing burden of aLRTD on healthcare systems and individual patients. An urgent public health need to improve the underlying health of patients with or at high-risk of respiratory disease remains. Measures such as acute respiratory hubs, pulmonary rehabilitation, effective smoking cessation and vaccination programmes with currently licensed influenza, SARS-CoV-2, pneumococcal, RSV and other vaccine against respiratory pathogens may help lessen the burden of respiratory disease [37], in line with recommendations from BTS, the Taskforce for Lung Health, and the NHS five-year plan.

## Data Availability

The data used in this study are sensitive and cannot be made publicly available without breaching patient confidentiality rules. Therefore, individual participant data and a data dictionary are not available to other researchers.

## ACKNOWLEDGEMENTS

We thank colleagues at the University of Bristol for their support with this study, including Rachel Davies, Paul Savage, Emma Foose, Susan Christie, Mark Mummé, Alison Horne, Mai Baquedano, and Adam Taylor. We would like to thank Stewart Robinson, David Clint and Henry Stuart and their teams for the support provided during this study. We would also like to acknowledge the research teams at North Bristol and University Hospitals of Bristol and Weston NHS Trusts for making this study possible, including Helen Lewis-White, Rebecca Smith, Rajeka Lazarus, Jane Blazeby, Diana Benton, and David Wynick. Finally, we would like to thank Aman Kaur-Singh and Kevin Sweetland for their support with this study.

## AUTHORSHIP STATEMENT

CH, RC, ML, EB, JS, CT, BG, AF, LD generated the research questions and analysis plans. The AvonCAP research team, MC, SM and CH were involved in data collection. CH, RC, LD and AF undertook data analysis. CH, RC, ML, EB, SMcG, MC, JS, JC, JO, CT, GE, NM, BG, LD, AF contributed to preparation of the manuscript and its revisions before publication. CH and AF provided oversight of the research.

## DATA SHARING

### The AvonCAP Research Group

Aaran Sinclair, Alessandra Mantini, Alexander Tremaine, Alison Horne, Amelia Langdon, Amy Taylor, Anabella Turner, Anna Jones, Anna Morley, Anya Mattocks, Archana Sudharsanan, Bethany Osborne, Brianna Dooley, Claire Mitchell, Emma Bridgeman, Emma Dickason-Palmer, Emma Scott, Ffion Davies, Fiona Perkins, Francesca Bayley, Francis Mensah, Gabriella Ruffino, Gabriella Valentine, Genevive Coulter, Grace Tilzey, Harriet Ibbotson, Jane Kinney, Johanna Kellett Wright, Joseph Cleere, Josephine Bonnici, Juan Garcia Tello Julia Brzezinska Julie Cloake, Katarina Milutinovic, Kate Helliker, Katie Maughan, Kazminder Fox, Konstantina Minou, Lana Ward, Laura Jaramillo, Leah Fleming, Leigh Morrison, Lily Smart, Lisa Grimmer, Louise Wright, Lucy Grimwood, Maddalena Bellavia, Maddie Clout, Mai Baquedano, Maise Borril, Maria Garcia Gonzalez, Marianne Vasquez, Martina Chmelarova, Matthew Jepson, Milo Jeenes-Flanagan, Natalie Chang, Nefeli Taravira, Niall Grace, Nicola Manning, Oliver Griffiths, Peter Sequenza, Pip Croxford, Rajeka Lazarus, Rebecca Abou Chullieh, Rebecca Clemence, Rhian Walters, Riley Cooper, Robert Graham, Robin Marlow, Robyn Heath, Rupert Antico, Sandi Nammuni Arachchge, Sarah Jenkins, Sean Robinson, Seevakumar Suppiah, Serena McGuinness, Siddiqa Uddin, Taslima Mona, Tawassal Riaz, Thomas Wright, Vicki Mackay, Zandile Maseko, Zoe Taylor, Zsolt Friedrich, Zsuzsa Szasz-Benczur

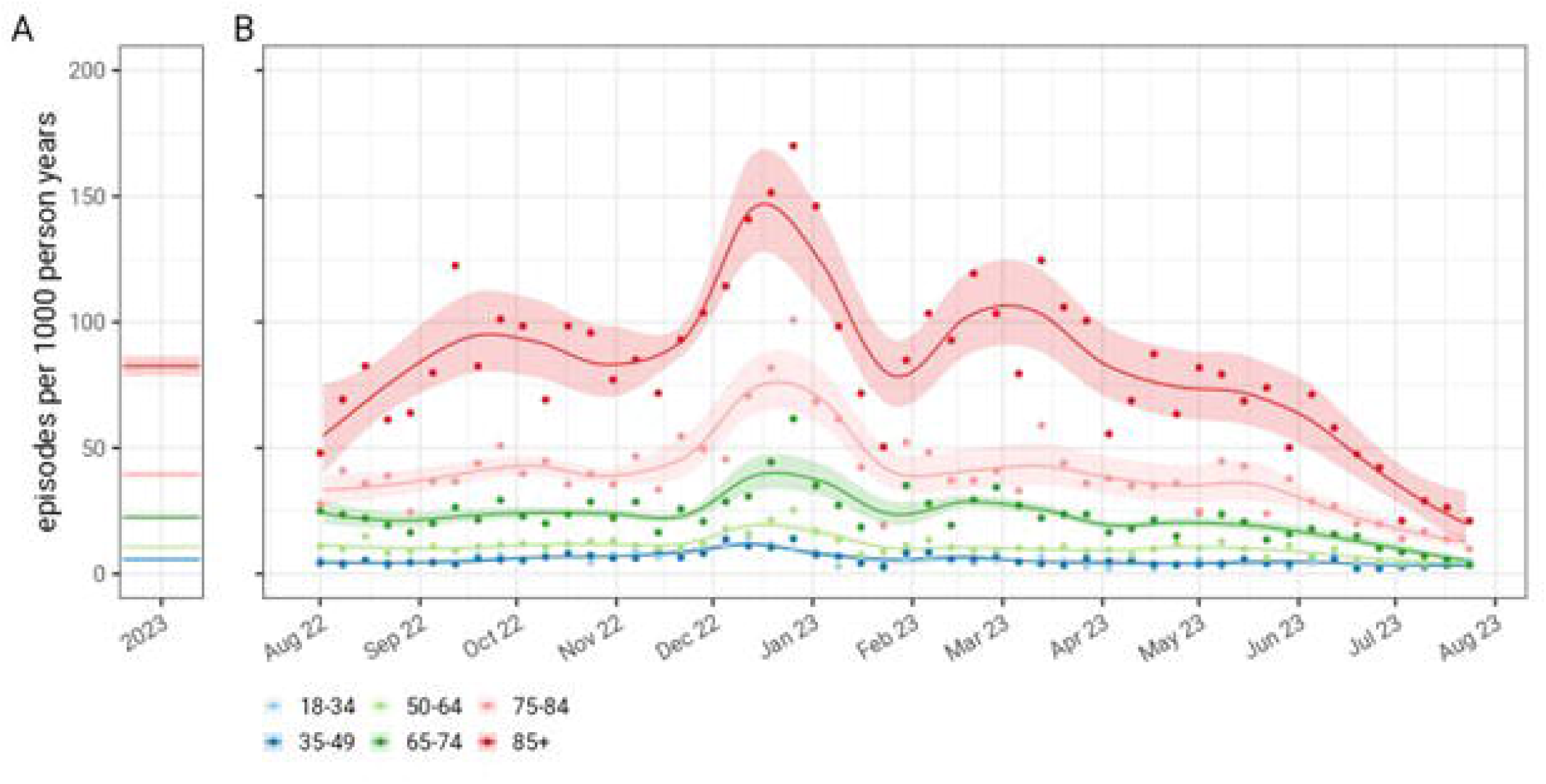

