## Supplementary Data for "Emergency department presentations of community-acquired lower respiratory tract disease in Bristol, UK: a prospective cohort study 2022-2023"

*Authors (need to be updated on Pfizer side)*

Dr Catherine Hyams MB PhD^1,2 *^, Dr Robert Challen MBBS PhD^3 *^, Dr Maria Lahuerta PhD MPH^4^, Dr Jo Southern PhD^5^, Ms Serena McGuinness MSc^1^, Ms Madeleine Clout MSc^2^, Dr Elizabeth Begier MD MPH^4^, Mr James Campling MSc^6^, Dr Jennifer Oliver PhD^1^, Dr Christian Theilacker^4^, Dr Gillian Ellsbury MD6, Prof Nick Maskell MD^2^, Dr Bradford Gessner PhD^4^, Dr Leon Danon PhD^3^, Professor Adam Finn PhD^1^ on behalf of the Avon CAP Research Group

^1^ Bristol Vaccine Centre, Population Health Sciences, University of Bristol, UK.

^2^ Academic Respiratory Unit, University of Bristol, UK.

^3^ Engineering Mathematics, University of Bristol, UK

^4^ Global Respiratory Vaccines, Medical & Scientific Affairs, Pfizer Vaccines, Collegeville, PA, USA

^5^ Evidence Generation, Pfizer, Collegeville, PA, USA

^6^ Vaccines Medical Affairs, Pfizer Ltd, Tadworth, UK, KT20 7NS.

^7^ Clinical Research and Imaging Centre, UHBW NHS Trust, Bristol, UK

* These authors contributed equally to the manuscript and should be considered co-first authors

* These authors contributed equally to the manuscript and should be considered co-first authors

*Corresponding Author:*

Catherine Hyams

Bristol Vaccine Centre, University of Bristol

39-41 St Michael’s Hill

Bristol BS2 8AE

United Kingdom

+44(0)117 342 0160

Keywords: pneumonia, lower respiratory tract infection

**Supplementary Table 1: Case Definitions**

| **Condition** | **Definition** | **Reference** |
| --- | --- | --- |
| Acute Lower Respiratory Tract Disease (aLRTD) | Acute lower respiratory tract disease (aLRTD) encompasses pneumonia, lower respiratory tract infection (LRTI), acute bronchitis, exacerbation of underlying respiratory disease including asthma and chronic obstructive pulmonary disease (COPD). Pneumothorax, pulmonary embolism, progression or new diagnosis of primary or secondary lung malignancy were excluded from aLRTD. | Hyams et al, BMJ Open 2022;12:e057464. doi: 10.1136/bmjopen-2021-057464BMJ Open paper |
| Pneumonia | Pneumonia was defined as infection affecting the airways (below the level of the larynx), with either (1) an acute illness with radiographic shadowing which was at least segmental or present in more than one lobe and was not known to be previously present or due to other causes, OR (2) in the absence of radiological investigation, clinical confirmation of pneumonic disease in the opinion of the treating physician | Lim WS, Baudouin SV, George RC, et al; Pneumonia Guidelines Committee of the BTS Standards of Care Committee. BTS guidelines for the management of community acquired pneumonia in adults: update 2009. Thorax. 2009 Oct;64 Suppl 3:iii1-55. |
| Non-Radiologically Proven Lower Respiratory Tract Infection (NP-LRTI) | An infection that affects the airways (below the level of the larynx) including the trachea and alveoli, with neither the presence of radiological change nor a clinical diagnosis of pneumonia from the treating physician, i.e. non-pneumonic infection in the lungs. | Anderson W, Winter J. Managing LRTI in adults in the community. Practitioner. 2009 Nov;253(1723):21-5, 2-3. PMID: 20043506. |
| Lung Abscess | A lung abscess was defined as a circumscribed area of pus or necrotic debris in lung parenchyma, which leads to a cavity with an air-fluid level inside the cavity | Bui JT, Schranz AJ, Strassle PD, Agala CB, Mody GN, Ikonomidis JS, Long JM. PLoS ONE. 2021 Sep 3; 16(9): e0256757 |
| Empyema | Empyema was defined as the presence of pus in the pleural space, and in this analysis we included complex parapneumonic effusion (CPE) in this category. Therefore, effusions with pleural fluid pH<7.2 and/or low glucose, elevated lactate dehydrogenase (LDH) and neutrophilia in addition to those with loculations due to infection were included in this category. | Roy B, Hui JS, Lee YC. Pleural fluid investigations for pleural infections. JLPM. 2021; 6 |
| Cardiac/Heart Failure (HF) | A clinical syndrome with symptoms and/or signs caused by a structural and/or functional cardiac abnormality and corroborated by elevated natriuretic peptide levels and or objective evidence of pulmonary or systemic congestion. | Bozkurt, Biykem et al.Universal Definition and Classification of Heart Failure. Journal of Cardiac Failure, Volume 27, Issue 4, 387 – 413. |
| Chronic Respiratory Disease Exacerbation (CRDE) | An acute (≤28 day) deterioration in respiratory function with the presence of signs/symptoms of aLRTD in a patient with a chronic respiratory disease, in line with the accepted definition of an acute exacerbation for the given chronic respiratory definition. |  |
| No evidence aLRTD | An acute (≤28 day) deterioration in respiratory function where there was neither a clinical nor microbiological diagnosis of infection. This includes non-infective exacerbations of chronic respiratory disease (e.g. asthma, COPD) and HF where there is no associated pulmonary infection component |  |

**Supplementary Table 2: Characteristics of patients seen in ED with aLRTD**

|  |  | All aLRTD | Seen in ED, but not admitted | Admitted from ED |  |
| --- | --- | --- | --- | --- | --- |
| Variable | Characteristic | Value (N=9452) | Value (N=2376) | Value (N=7076) | P value |
| Age | Median [IQR] | 69.2 [48.6—81.1] | 43.4 [29.4—62.3] | 74 [59.8—83.5] | <0.001 † |
| Age Category | 18-34 % (n) | 14.2% (1339) | 37.0% (879) | 6.5% (460) | <0.001 †† |
|  | 35-49 % (n) | 11.9% (1123) | 21.7% (516) | 8.6% (607) |  |
|  | 50-64 % (n) | 17.8% (1681) | 19.3% (459) | 17.3% (1222) |  |
|  | 65-74 % (n) | 17.4% (1640) | 8.6% (205) | 20.3% (1435) |  |
|  | 75-84 % (n) | 21.6% (2046) | 8.7% (206) | 26.0% (1840) |  |
|  | 85+ % (n) | 17.2% (1623) | 4.7% (111) | 21.4% (1512) |  |
| Age Eligible for PneumoVax | 18-64 % (n) | 43.8% (4143) | 78.0% (1854) | 32.3% (2289) | <0.001 †† |
|  | 65+ % (n) | 56.2% (5309) | 22.0% (522) | 67.7% (4787) |  |
| Gender | Female % (n) | 52.1% (4926) | 54.8% (1301) | 51.2% (3625) | 0.003 †† |
| Care Home Resident | no % (n) | 93.0% (8787) | 97.5% (2316) | 91.4% (6471) | <0.001 †† |
|  | yes % (n) | 7.0% (662) | 2.4% (58) | 8.5% (604) |  |
|  | <missing> % (n) | 0.0% (3) | 0.1% (2) | 0.0% (1) |  |
| Smoker | Non-smoker % (n) | 38.8% (3665) | 51.1% (1214) | 34.6% (2451) | <0.001 †† |
|  | Current % (n) | 15.1% (1428) | 14.6% (348) | 15.3% (1080) |  |
|  | Ex-smoker % (n) | 40.1% (3792) | 27.3% (649) | 44.4% (3143) |  |
|  | Unknown % (n) | 6.0% (567) | 6.9% (165) | 5.7% (402) |  |
| IMD (decile) | Median [IQR] | 5 [3—8] | 5 [3—8] | 5 [3—8] | — ††† |
| Alcohol Abuse | yes % (n) | 5.3% (502) | 4.3% (101) | 5.7% (401) | 0.007 †† |
| aLTRD presentation | Pneumonia % (n) | 38.8% (3663) | 17.0% (405) | 46.0% (3258) | <0.001 †† |
|  | NP-LRTI % (n) | 44.1% (4167) | 63.3% (1504) | 37.6% (2663) |  |
|  | No evidence LRTI % (n) | 17.2% (1622) | 19.7% (467) | 16.3% (1155) |  |
| Heart Failure | yes % (n) | 14.6% (1376) | 3.0% (72) | 18.4% (1304) | <0.001 †† |
| Exacerbation Of Chronic Respiratory Disease | yes % (n) | 48.1% (4551) | 45.4% (1079) | 49.1% (3472) | 0.002 †† |
| CCI Category | None (0) % (n) | 21.9% (2066) | 52.9% (1257) | 11.4% (809) | <0.001 †† |
|  | Mild (1-2) % (n) | 14.1% (1337) | 19.5% (463) | 12.4% (874) |  |
|  | Moderate (3-4) % (n) | 25.2% (2381) | 14.0% (332) | 29.0% (2049) |  |
|  | Severe (5+) % (n) | 38.8% (3668) | 13.6% (324) | 47.3% (3344) |  |
| CURB 65 Category | 0-1 (Mild) % (n) | 80.1% (7575) | 96.8% (2299) | 74.6% (5276) | <0.001 †† |
|  | 2 (Moderate) % (n) | 17.8% (1680) | 3.1% (74) | 22.7% (1606) |  |
|  | 3-5 (Severe) % (n) | 2.1% (197) | 0.1% (3) | 2.7% (194) |  |
| Rockwood Category | Independent (0-4) % (n) | 53.4% (5044) | 46.0% (1092) | 55.9% (3952) | <0.001 †† |
|  | Frail (5-9) % (n) | 46.6% (4407) | 54.0% (1284) | 44.1% (3123) |  |
|  | <missing> % (n) | 0.0% (1) | 0.0% (0) | 0.0% (1) |  |
| Pneumonia Severity Index Class | Class I % (n) | 18.8% (1779) | 44.9% (1066) | 10.1% (713) | <0.001 †† |
|  | Class II % (n) | 22.5% (2127) | 34.5% (820) | 18.5% (1307) |  |
|  | Class III % (n) | 19.5% (1846) | 10.9% (259) | 22.4% (1587) |  |
|  | Class IV % (n) | 28.5% (2693) | 8.1% (193) | 35.3% (2500) |  |
|  | Class V % (n) | 10.7% (1007) | 1.6% (38) | 13.7% (969) |  |
| NEWS2 Score | Median [IQR] | 3 [1—5] | 1 [0—3] | 3 [1—6] | <0.001 †††† |
| COPD | yes % (n) | 27.4% (2590) | 13.7% (325) | 32.0% (2265) | <0.001 †† |
| Asthma | yes % (n) | 19.5% (1839) | 30.5% (724) | 15.8% (1115) | <0.001 †† |
| Bronchiectasis | yes % (n) | 3.7% (347) | 1.5% (36) | 4.4% (311) | <0.001 †† |
| Interstitial Lung Dx | yes % (n) | 2.0% (193) | 0.8% (18) | 2.5% (175) | <0.001 †† |
| Any Chronic Lung Disease | yes % (n) | 48.3% (4563) | 43.8% (1040) | 49.8% (3523) | <0.001 †† |
| IHD | yes % (n) | 11.5% (1084) | 5.5% (131) | 13.5% (953) | <0.001 †† |
| Hypertension | yes % (n) | 14.5% (1366) | 5.5% (130) | 17.5% (1236) | <0.001 †† |
| Any Chronic Heart Disease | yes % (n) | 35.4% (3349) | 14.7% (350) | 42.4% (2999) | <0.001 †† |
| Immunodeficiency | yes % (n) | 0.4% (38) | 0.2% (4) | 0.5% (34) | 0.039 †† |
| On Immunosuppression | no % (n) | 84.2% (7961) | 83.5% (1983) | 84.5% (5978) | <0.001 †† |
|  | yes % (n) | 15.5% (1469) | 15.6% (371) | 15.5% (1098) |  |
|  | <missing> % (n) | 0.2% (22) | 0.9% (22) | 0.0% (0) |  |
| Any Immune Compromise* | yes % (n) | 15.8% (1495) | 15.7% (373) | 15.9% (1122) | 0.87 †† |
| Diabetes Type | None % (n) | 81.8% (7735) | 90.9% (2160) | 78.8% (5575) | <0.001 †† |
|  | Type 1 % (n) | 1.0% (93) | 0.5% (11) | 1.2% (82) |  |
|  | Type 2 % (n) | 17.2% (1624) | 8.6% (205) | 20.1% (1419) |  |
| CKD | None % (n) | 77.6% (7337) | 93.0% (2209) | 72.5% (5128) | <0.001 †† |
|  | Mild (CKD 1-3) % (n) | 19.1% (1803) | 6.4% (151) | 23.3% (1652) |  |
|  | Moderate or Severe CKD (CKD 4+) % (n) | 3.3% (312) | 0.7% (16) | 4.2% (296) |  |
| CVA/TIA | yes % (n) | 9.3% (880) | 3.1% (73) | 11.4% (807) | <0.001 †† |
| Any Cancer Present | yes % (n) | 9.8% (926) | 3.9% (92) | 11.8% (834) | <0.001 †† |
| Pneumovax | Not received % (n) | 54.0% (5104) | 73.7% (1752) | 47.4% (3352) | <0.001 †† |
|  | PPV23 % (n) | 37.4% (3537) | 14.7% (349) | 45.1% (3188) |  |
|  | PCV13 % (n) | 0.9% (88) | 0.9% (21) | 0.9% (67) |  |
|  | Unknown % (n) | 7.6% (722) | 10.6% (253) | 6.6% (469) |  |
|  | <missing> % (n) | 0.0% (1) | 0.0% (1) | 0.0% (0) |  |
| Influenza Vaccination | Not received % (n) | 36.5% (3449) | 58.0% (1378) | 29.3% (2071) | <0.001 †† |
|  | Received % (n) | 60.8% (5745) | 37.3% (887) | 68.7% (4858) |  |
|  | Unknown % (n) | 2.7% (258) | 4.7% (111) | 2.1% (147) |  |
| COVID-19 Vaccination | Not received % (n)  Received ≥2 doses % (n) | 80.0% (7565)  20.0% (1887) | 57.1% (1356)  43.0% (1020) | 87.7% (6209)  12.3% (867) | <0.001 †† |
| Dyspnoea | yes % (n) | 73.0% (6902) | 68.6% (1630) | 74.5% (5272) | <0.001 †† |
| Cough | yes % (n) | 65.0% (6142) | 72.6% (1725) | 62.4% (4417) | <0.001 †† |
| Fever | yes % (n) | 23.6% (2230) | 26.3% (624) | 22.7% (1606) | <0.001 †† |
|  | no % (n) | 76.3% (7209) | 73.3% (1742) | 77.3% (5467) |  |
|  | unknown % (n) | 0.1% (12) | 0.4% (10) | 0.0% (2) |  |
|  | <missing> % (n) | 0.0% (1) | 0.0% (0) | 0.0% (1) |  |
| Confusion | no % (n) | 93.6% (8851) | 98.9% (2349) | 91.9% (6502) | <0.001 †† |
|  | yes % (n) | 6.3% (600) | 1.1% (27) | 8.1% (573) |  |
|  | <missing> % (n) | 0.0% (1) | 0.0% (0) | 0.0% (1) |  |
| Oxygen Requirement | 21 % (n) | 72.6% (6862) | 94.2% (2238) | 65.3% (4624) | <0.001 †† |
|  | 22‒27 % (n) | 4.2% (396) | 0.8% (20) | 5.3% (376) |  |
|  | 28‒34 % (n) | 11.4% (1075) | 1.6% (37) | 14.7% (1038) |  |
|  | 35‒49 % (n) | 5.6% (527) | 0.3% (7) | 7.3% (520) |  |
|  | 50‒100 % (n) | 5.4% (511) | 0.5% (13) | 7.0% (498) |  |
|  | <missing> % (n) | 0.9% (81) | 2.6% (61) | 0.3% (20) |  |
| Oximetry | Median [IQR] | 96 [94—98] | 97 [96—99] | 96 [93—97] | — ††† |
| Heart Rate | Median [IQR] | 90 [78—105] | 90 [78—102] | 90 [78—105] | — ††† |
| Respiratory Rate | Median [IQR] | 20 [18—23] | 19 [17—20] | 20 [18—24] | — ††† |
| Systolic BP | Median [IQR] | 130 [115—146] | 130 [119—145] | 130 [114—146] | — ††† |
| Diastolic BP | Median [IQR] | 76 [67—86] | 80 [73—89] | 75 [65—85] | — ††† |
| White Cell Count | Median [IQR] | 10 [7.44—13.6] | 8.94 [6.86—11.5] | 10.4 [7.6—14.2] | — ††† |
| CRP | Median [IQR] | 31 [9—93] | 14 [4—44] | 38 [11—109] | — ††† |
| Urea | Median [IQR] | 5.8 [4.1—8.7] | 4.4 [3.4—5.7] | 6.4 [4.4—9.7] | — ††† |
| Radiology Investigation | performed % (n) | 72.3% (6835) | 61.3% (1457) | 76.0% (5378) | <0.001 †† |
| Virology Testing | performed % (n) | 57.1% (5394) | 42.4% (1007) | 62.0% (4387) | <0.001 †† |
| Microbiology Testing | performed % (n) | 23.2% (2192) | 6.1% (146) | 28.9% (2046) | <0.001 †† |
| †, 2 sample Kolmogorov-Smirnov test (continuous); ††, Fisher's exact test (categorical); †††, Not calculated due to missing values (continuous); ††††, 2 sample Wilcoxon Rank Sum test (continuous)  Normal distributions determined by the Anderson-Darling test (P>0.005)  An adjusted P value of 0.00102 may be considered significant. | | | | | |

aLRTD, acute lower respiratory tract disease; BP, blood pressure; CCI, Charlson Comorbidity; CKD, chronic kidney disease; CVA, cerebrovascular disease; COPD, chronic obstructive pulmonary disease; CRP, C-reactive protein; ED, emergency department; IHD, ischaemic heart disease; IMD, Index of multiple deprivation (decile); NEWS-2, new early warning system; TIA, transient ischaemic attack

* Participants were classified as high risk (i.e., immunocompromised) if they had chronic kidney disease, organ transplantation, immunodeficiency, haematological or solid malignancy, acquired immunodeficiency syndrome (AIDS), human immunodeficiency virus (HIV) infection, or were treated with immunosuppressive drug therapy

**Supplementary Table 3: Frequency of missing data stratified by admission to hospital.**

Percentages reflect the rate of missing values, for each variable. P-values test the null hypothesis that the data is missing at random between the two groups.

|  | Seen in ED, but not admitted | Admitted from ED |  |
| --- | --- | --- | --- |
| variable | missing % (N) | missing % (N) | P value |
| Age | 0.0% (0/2376) | 0.0% (0/7076) | 1 |
| Age Category | 0.0% (0/2376) | 0.0% (0/7076) | 1 |
| Age Eligible for PneumoVax | 0.0% (0/2376) | 0.0% (0/7076) | 1 |
| Gender | 0.0% (0/2376) | 0.0% (0/7076) | 1 |
| Care Home Resident | 0.1% (2/2376) | 0.0% (1/7076) | 0.16 |
| Smoker | 0.0% (0/2376) | 0.0% (0/7076) | 1 |
| IMD (decile) | 3.5% (84/2376) | 0.9% (62/7076) | <0.001 |
| Alcohol Abuse | 0.0% (0/2376) | 0.0% (0/7076) | 1 |
| aLTRD presentation | 0.0% (0/2376) | 0.0% (0/7076) | 1 |
| Heart Failure | 0.0% (0/2376) | 0.0% (0/7076) | 1 |
| Exacerbation Of Chronic Respiratory Disease | 0.0% (0/2376) | 0.0% (0/7076) | 1 |
| CCI Category | 0.0% (0/2376) | 0.0% (0/7076) | 1 |
| CURB 65 Category | 0.0% (0/2376) | 0.0% (0/7076) | 1 |
| Rockwood Category | 0.0% (0/2376) | 0.0% (1/7076) | 1 |
| Pneumonia Severity Index Class | 0.0% (0/2376) | 0.0% (0/7076) | 1 |
| NEWS2 Score | 0.0% (0/2376) | 0.0% (0/7076) | 1 |
| COPD | 0.0% (0/2376) | 0.0% (0/7076) | 1 |
| Asthma | 0.0% (0/2376) | 0.0% (0/7076) | 1 |
| Bronchiectasis | 0.0% (0/2376) | 0.0% (0/7076) | 1 |
| Interstitial Lung Dx | 0.0% (0/2376) | 0.0% (0/7076) | 1 |
| Any Chronic Lung Disease | 0.0% (0/2376) | 0.0% (0/7076) | 1 |
| IHD | 0.0% (0/2376) | 0.0% (0/7076) | 1 |
| Hypertension | 0.0% (0/2376) | 0.0% (0/7076) | 1 |
| Any Chronic Heart Disease | 0.0% (0/2376) | 0.0% (0/7076) | 1 |
| Immunodeficiency | 0.0% (0/2376) | 0.0% (0/7076) | 1 |
| On Immunosuppression | 0.9% (22/2376) | 0.0% (0/7076) | <0.001 |
| Any Immune Compromise | 0.0% (0/2376) | 0.0% (0/7076) | 1 |
| Diabetes Type | 0.0% (0/2376) | 0.0% (0/7076) | 1 |
| CKD | 0.0% (0/2376) | 0.0% (0/7076) | 1 |
| CVA/TIA | 0.0% (0/2376) | 0.0% (0/7076) | 1 |
| Any Cancer Present | 0.0% (0/2376) | 0.0% (0/7076) | 1 |
| Pneumovax | 0.0% (1/2376) | 0.0% (0/7076) | 0.25 |
| Influenza Vaccination | 0.0% (0/2376) | 0.0% (0/7076) | 1 |
| Dyspnoea | 0.0% (0/2376) | 0.0% (0/7076) | 1 |
| Cough | 0.0% (0/2376) | 0.0% (0/7076) | 1 |
| Fever | 0.0% (0/2376) | 0.0% (1/7076) | 1 |
| Confusion | 0.0% (0/2376) | 0.0% (1/7076) | 1 |
| Oxygen Requirement (FiO_2_) | 0.0% (0/2376) | 0.0% (0/7076) | 1 |
| Oximetry | 2.6% (62/2376) | 0.3% (24/7076) | <0.001 |
| Heart Rate | 3.1% (74/2376) | 0.3% (23/7076) | <0.001 |
| Respiratory Rate | 4.0% (95/2376) | 0.4% (26/7076) | <0.001 |
| Systolic BP | 4.1% (98/2376) | 0.4% (25/7076) | <0.001 |
| Diastolic BP | 4.2% (99/2376) | 0.4% (29/7076) | <0.001 |
| White Cell Count | 22.1% (526/2376) | 0.9% (66/7076) | <0.001 |
| CRP | 31.4% (747/2376) | 2.5% (175/7076) | <0.001 |
| Urea | 22.4% (532/2376) | 1.0% (68/7076) | <0.001 |
| Radiology Investigation | 0.0% (0/2376) | 0.0% (0/7076) | 1 |
| Virology Testing | 0.0% (0/2376) | 0.0% (0/7076) | 1 |
| Microbiology Testing | 0.0% (0/2376) | 0.0% (0/7076) | 1 |
| Data is missing not at random (compared to Episode Type) at a p-value<0.001 (0.05 over 49 comparisons) for variables IMD (decile), On Immunosuppression, Oximetry, Heart Rate, Respiratory Rate, Systolic BP, Diastolic BP, White Cell Count, CRP, Urea. | | | |

aLRTD, acute lower respiratory tract disease; BP, blood pressure; CCI, Charlson Comorbidity; CKD, chronic kidney disease; CVA, cerebrovascular disease; COPD, chronic obstructive pulmonary disease; CRP, C-reactive protein; ED, emergency department; FiO_2_, fraction of inspired oxygen; IHD, ischaemic heart disease; IMD, Index of multiple deprivation (decile); NEWS-2, new early warning system; TIA, transient ischaemic attack

**Supplementary Table 4: Univariate and minimally age adjusted logistic regression models**

Univariate Logistic regression models based on a-priori covariates, for which there was no missing data are shown in the left hand column. Age is the major determinant of admission. The same covariates adjusted for categorical age are in the right-hand column. Data items with major correlations with age, resulting from for example, vaccination policy, which may be acting as a proxy for age in the models, were reviewed for suitability for inclusion in the next step of analysis. Associations which became non-significant when adjusted for age were also excluded at this stage.

|  |  | Hospital Admission (a. Univariate) (N=9452) | | Hospital Admission (b. Age adjusted) (N=9452) | |
| --- | --- | --- | --- | --- | --- |
| Characteristic | Subgroup | OR [95% CI] | P value | OR [95% CI] | P value |
| Age Category | 18-34 | ref | <0.001 | ref | <0.001 |
|  | 35-49 | 2.2 [1.9 – 2.6] |  | — |  |
|  | 50-64 | 5.1 [4.4 – 5.9] |  | — |  |
|  | 65-74 | 13 [11 – 16] |  | — |  |
|  | 75-84 | 17 [14 – 20] |  | — |  |
|  | 85+ | 26 [21 – 33] |  | — |  |
| Gender | Female | ref | 0.003 | ref | 0.423 |
|  | Male | 1.2 [1 – 1.3] |  | 1 [0.94 – 1.2] |  |
| Care Home Resident | no | ref | <0.001 | ref | <0.001 |
|  | yes | 3.7 [2.8 – 4.9] |  | 1.7 [1.3 – 2.3] |  |
| Smoker | Non-smoker | ref | <0.001 | ref | <0.001 |
|  | Current | 1.5 [1.3 – 1.8] |  | 1.7 [1.4 – 2] |  |
|  | Ex-smoker | 2.4 [2.2 – 2.7] |  | 1.4 [1.2 – 1.5] |  |
|  | Unknown | 1.2 [0.99 – 1.5] |  | 1.3 [1 – 1.6] |  |
| IMD (decile) |  | 1 [0.99 – 1] | 0.342 | 0.96 [0.95 – 0.98] | <0.001 |
| CCI Category | None (0) | ref | <0.001 | ref | <0.001 |
|  | Mild (1-2) | 2.9 [2.5 – 3.4] |  | 2 [1.6 – 2.6] |  |
|  | Moderate (3-4) | 9.6 [8.3 – 11] |  | 4.3 [3.2 – 5.8] |  |
|  | Severe (5+) | 16 [14 – 19] |  | 5.8 [4.2 – 8] |  |
| CURB 65 Category | 0-1 (Mild) | ref | <0.001 | ref | <0.001 |
|  | 2 (Moderate) | 9.5 [7.5 – 12] |  | 3.4 [2.6 – 4.3] |  |
|  | 3-5 (Severe) | 28 [9 – >50] |  | 9.4 [3 – 30] |  |
| aLTRD presentation | Pneumonia | ref | <0.001 | ref | <0.001 |
|  | NP-LRTI | 0.22 [0.2 – 0.25] |  | 0.32 [0.28 – 0.36] |  |
|  | No evidence LRTI | 0.31 [0.26 – 0.36] |  | 0.39 [0.33 – 0.46] |  |
| Heart Failure | no | ref | <0.001 | ref | <0.001 |
|  | yes | 7.2 [5.7 – 9.2] |  | 3.4 [2.7 – 4.4] |  |
| Exacerbation Of Chronic Respiratory Disease | no | ref | 0.002 | ref | 0.056 |
|  | yes | 1.2 [1.1 – 1.3] |  | 1.1 [1 – 1.2] |  |
| Any Chronic Heart Disease | no | ref | <0.001 | ref | <0.001 |
|  | yes | 4.3 [3.8 – 4.8] |  | 1.6 [1.4 – 1.9] |  |
| Diabetes Type | None | ref | <0.001 | ref | <0.001 |
|  | Type 1 | 2.9 [1.5 – 5.4] |  | 3.9 [2 – 7.6] |  |
|  | Type 2 | 2.7 [2.3 – 3.1] |  | 1.4 [1.2 – 1.7] |  |
| Pneumovax | Unknown | 0.97 [0.82 – 1.1] | <0.001 | 0.81 [0.68 – 0.98] | <0.001 |
|  | Not received | ref |  | ref |  |
|  | PPV23 | 4.8 [4.2 – 5.4] |  | 1.7 [1.5 – 2] |  |
|  | PCV13 | 1.7 [1 – 2.7] |  | 0.77 [0.45 – 1.3] |  |
| Influenza Vaccination | Unknown | 0.88 [0.68 – 1.1] | <0.001 | 0.92 [0.69 – 1.2] | <0.001 |
|  | Not received | ref |  | ref |  |
|  | Received | 3.6 [3.3 – 4] |  | 1.3 [1.2 – 1.5] |  |
| Dyspnoea | no | ref | <0.001 | ref | <0.001 |
|  | yes | 1.3 [1.2 – 1.5] |  | 1.3 [1.2 – 1.5] |  |
| Cough | no | ref | <0.001 | ref | <0.001 |
|  | yes | 0.63 [0.57 – 0.69] |  | 0.78 [0.69 – 0.87] |  |
| Fever | no | ref | <0.001 | ref | <0.001 |
|  | yes | 0.82 [0.74 – 0.91] |  | 1.2 [1 – 1.3] |  |
|  | unknown | 0.064 [<0.02 – 0.29] |  | 0.12 [0.022 – 0.63] |  |
| Confusion | no | ref | <0.001 | ref | <0.001 |
|  | yes | 7.7 [5.2 – 11] |  | 3 [2 – 4.4] |  |
| Oxygen Requirement | unknown | 0.15 [0.09 – 0.25] | <0.001 | 0.18 [0.1 – 0.33] | <0.001 |
|  | 21 | ref |  | ref |  |
|  | 22‒27 | 9.1 [5.8 – 14] |  | 5.4 [3.4 – 8.7] |  |
|  | 28‒34 | 14 [9.7 – 19] |  | 8.8 [6.3 – 12] |  |
|  | 35‒100 | 25 [16 – 38] |  | 18 [12 – 29] |  |
| P values calculated using Likelihood ratio test (II) | | | | | |

CCI, Charlson comorbidity index; CI, confidence interval; FiO_2_, fraction of inspired oxygen; IMD, index of multiple deprivation; LRTI, lower respiratory tract infection; NP-LRTI, non-pneumonic lower respiratory tract infection; N, number; OR, odds ratio; PCV-13, pneumococcal conjugate vaccine (13-valent); PPV23, pneumococcal polysaccharide vaccine (23-valent); y, years.

**Supplementary Table 5: Fully adjusted logistic regression models involving non interacting terms**

This fully adjusted multiple logistic regression model is based on a restricted, but still comprehensive set of covariates that were not seen to be interacting heavily with age to identify variables with residual significant impact on the outcome. From these remaining variables a parsimonious model was constructed which is presented in the main paper.

|  |  | Hospital Admission (N=9305) | |
| --- | --- | --- | --- |
| Characteristic | Subgroup | OR [95% CI] | P value |
| Age Category | 18-34 | ref | <0.001 |
|  | 35-49 | 1.6 [1.3 – 1.9] |  |
|  | 50-64 | 1.6 [1.2 – 2.2] |  |
|  | 65-74 | 2.7 [1.9 – 3.9] |  |
|  | 75-84 | 2.9 [2 – 4.3] |  |
|  | 85+ | 4.1 [2.7 – 6.2] |  |
| Smoker | Non-smoker | ref | <0.001 |
|  | Current | 1.5 [1.3 – 1.8] |  |
|  | Ex-smoker | 1.3 [1.1 – 1.5] |  |
|  | Unknown | 1.4 [1.1 – 1.7] |  |
| IMD (decile) |  | 0.98 [0.96 – 1] | 0.100 |
| CCI Category | None (0) | ref | <0.001 |
|  | Mild (1-2) | 1.4 [1.1 – 1.8] |  |
|  | Moderate (3-4) | 2.5 [1.8 – 3.6] |  |
|  | Severe (5+) | 2.6 [1.8 – 3.8] |  |
| aLTRD presentation | Pneumonia | ref | <0.001 |
|  | NP-LRTI | 0.41 [0.36 – 0.47] |  |
|  | No evidence LRTI | 0.38 [0.32 – 0.46] |  |
| Heart Failure | no | ref | <0.001 |
|  | yes | 2.7 [2.1 – 3.6] |  |
| Exacerbation Of Chronic Respiratory Disease | no | ref | 0.678 |
|  | yes | 1 [0.9 – 1.2] |  |
| Any Chronic Heart Disease | no | ref | 0.003 |
|  | yes | 1.3 [1.1 – 1.5] |  |
| Diabetes Type | None | ref | 0.029 |
|  | Type 1 | 2.4 [1.2 – 4.8] |  |
|  | Type 2 | 1 [0.83 – 1.2] |  |
| Dyspnoea | no | ref | 0.408 |
|  | yes | 1.1 [0.93 – 1.2] |  |
| Cough | no | ref | <0.001 |
|  | yes | 0.69 [0.6 – 0.78] |  |
| Fever | no | ref | 0.102 |
|  | yes | 1.1 [0.99 – 1.3] |  |
|  | unknown | 0.35 [0.043 – 2.8] |  |
| Confusion | no | ref | <0.001 |
|  | yes | 2.5 [1.7 – 3.8] |  |
| Oxygen Requirement | unknown | 0.22 [0.12 – 0.4] | <0.001 |
|  | 21 | ref |  |
|  | 22‒27 | 4.3 [2.7 – 6.9] |  |
|  | 28‒34 | 7.5 [5.3 – 11] |  |
|  | 35‒100 | 13 [8.5 – 21] |  |
| P values calculated using Likelihood ratio test (II) | | | |

**Supplementary Figure 1: Age adjusted and fully adjusted logistic regression models**

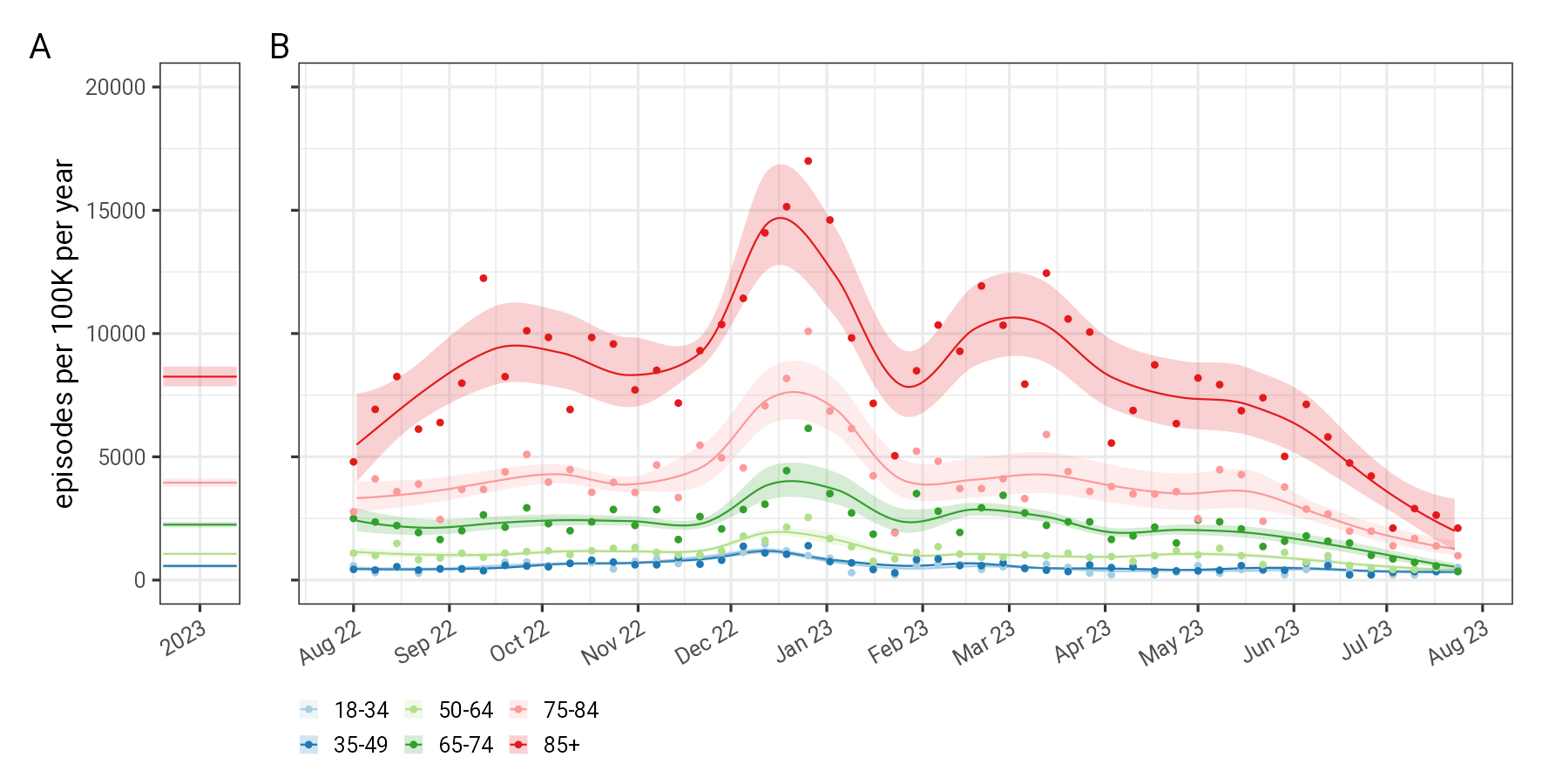

**Supplementary Table 6: Combined incidence of emergency department episodes for aLRTD per 100K persons per year**

Incidence by 100,000 persons per year based on the estimated population of the catchment area of the AvonCAP study hospitals are presented rounded to nearest 10 due to the sample size.

|  |  | All aLTRD | Seen in ED, but not admitted | | | | Admitted from ED | | | |
| --- | --- | --- | --- | --- | --- | --- | --- | --- | --- | --- |
|  |  | All cause | All cause | Pneumonia | NP-LRTI | No evidence LRTI | All cause | Pneumonia | NP-LRTI | No evidence LRTI |
| Age Group | Mid-year population | (N=9452)  per 100K per yr | (N=2376)  per 100K per yr | (N=405)  per 100K per yr | (N=1504)  per 100K per yr | (N=467)  per 100K per yr | (N=7076)  per 100K per yr | (N=3258)  per 100K per yr | (N=2663)  per 100K per yr | (N=1155)  per 100K per yr |
| All adults | 739651 | 1280 | 320 | 50 | 200 | 60 | 960 | 440 | 360 | 160 |
| 18-34 | 242415 | 550 | 360 | 30 | 260 | 70 | 190 | 60 | 100 | 30 |
| 35-49 | 194783 | 580 | 260 | 50 | 180 | 40 | 310 | 120 | 130 | 60 |
| 50-64 | 158100 | 1060 | 290 | 70 | 160 | 70 | 770 | 330 | 300 | 150 |
| 65-74 | 72929 | 2250 | 280 | 70 | 140 | 70 | 1970 | 910 | 730 | 340 |
| 75-84 | 51784 | 3950 | 400 | 100 | 220 | 80 | 3550 | 1770 | 1250 | 530 |
| 85+ | 19666 | 8250 | 560 | 200 | 250 | 110 | 7690 | 3920 | 2670 | 1100 |
| 18-64 | 595278 | 700 | 310 | 40 | 210 | 60 | 380 | 150 | 160 | 70 |
| 65+ | 144371 | 3680 | 360 | 100 | 180 | 80 | 3320 | 1630 | 1180 | 510 |

**Supplementary Table 7: Sensitivity analysis combined incidence of emergency department episodes for aLRTD per 1000 persons years by catchment area estimation methods.**

Larger denominators resulting in smaller incidence estimates and the largest variation is between the different estimates of the 75-84 and 85+ age groups. The “Campling 2019” model is a proportional flow model mapping aLRTD admissions to GP practices in the Bristol, Somerset, and South Gloucester Integrated Care Board, and is presented in the main paper. The “Challen 2019” model is a supply demand weighted label propagation model, and the “OHID (exc Weston) 2019: Emergency” model is another proportional flow model, mapping emergency admissions to middle super output areas, as detailed in Supplementary 2.

|  |  |  | All aLTRD | Seen in ED, but not admitted | | | | Admitted from ED | | | |
| --- | --- | --- | --- | --- | --- | --- | --- | --- | --- | --- | --- |
|  |  |  | All cause | All cause | Pneumonia | NP-LRTI | No evidence LRTI | All cause | Pneumonia | NP-LRTI | No evidence LRTI |
| Method | Age Group | Mid-year population | (N=9452)  per 1000 person yr | (N=2376)  per 1000 person yr | (N=405)  per 1000 person yr | (N=1504)  per 1000 person yr | (N=467)  per 1000 person yr | (N=7076)  per 1000 person yr | (N=3258)  per 1000 person yr | (N=2663)  per 1000 person yr | (N=1155)  per 1000 person yr |
| Campling 2019 | All adults | 739651 | 12.8 | 3.2 | 0.5 | 2.0 | 0.6 | 9.6 | 4.4 | 3.6 | 1.6 |
|  | 18-34 | 242415 | 5.5 | 3.6 | 0.3 | 2.6 | 0.7 | 1.9 | 0.6 | 1.0 | 0.3 |
|  | 35-49 | 194783 | 5.8 | 2.6 | 0.5 | 1.8 | 0.4 | 3.1 | 1.2 | 1.3 | 0.6 |
|  | 50-64 | 158100 | 10.6 | 2.9 | 0.7 | 1.6 | 0.7 | 7.7 | 3.3 | 3.0 | 1.5 |
|  | 65-74 | 72929 | 22.5 | 2.8 | 0.7 | 1.4 | 0.7 | 19.7 | 9.1 | 7.3 | 3.4 |
|  | 75-84 | 51784 | 39.5 | 4.0 | 1.0 | 2.2 | 0.8 | 35.5 | 17.7 | 12.5 | 5.3 |
|  | 85+ | 19666 | 82.5 | 5.6 | 2.0 | 2.5 | 1.1 | 76.9 | 39.2 | 26.7 | 11.0 |
|  | 18-64 | 595278 | 7.0 | 3.1 | 0.4 | 2.1 | 0.6 | 3.8 | 1.5 | 1.6 | 0.7 |
|  | 65+ | 144371 | 36.8 | 3.6 | 1.0 | 1.8 | 0.8 | 33.2 | 16.3 | 11.8 | 5.1 |
| Challen 2019 | All adults | 739980 | 12.8 | 3.2 | 0.5 | 2.0 | 0.6 | 9.6 | 4.4 | 3.6 | 1.6 |
|  | 18-34 | 251310 | 5.3 | 3.5 | 0.3 | 2.5 | 0.7 | 1.8 | 0.6 | 1.0 | 0.3 |
|  | 35-49 | 179757 | 6.2 | 2.9 | 0.5 | 1.9 | 0.4 | 3.4 | 1.3 | 1.4 | 0.6 |
|  | 50-64 | 156284 | 10.8 | 2.9 | 0.7 | 1.6 | 0.7 | 7.8 | 3.4 | 3.0 | 1.5 |
|  | 65-74 | 77349 | 21.2 | 2.7 | 0.7 | 1.3 | 0.7 | 18.6 | 8.5 | 6.8 | 3.2 |
|  | 75-84 | 54102 | 37.8 | 3.8 | 1.0 | 2.1 | 0.8 | 34.0 | 17.0 | 11.9 | 5.1 |
|  | 85+ | 21200 | 76.6 | 5.2 | 1.9 | 2.4 | 1.0 | 71.3 | 36.4 | 24.8 | 10.2 |
|  | 18-64 | 587335 | 7.1 | 3.2 | 0.4 | 2.1 | 0.6 | 3.9 | 1.5 | 1.6 | 0.7 |
|  | 65+ | 152643 | 34.8 | 3.4 | 0.9 | 1.7 | 0.8 | 31.4 | 15.4 | 11.1 | 4.8 |
| OHID (exc Weston) 2019: Emergency | All adults | 847604 | 11.2 | 2.8 | 0.5 | 1.8 | 0.6 | 8.3 | 3.8 | 3.1 | 1.4 |
|  | 18-34 | 279651 | 4.8 | 3.1 | 0.2 | 2.3 | 0.6 | 1.6 | 0.5 | 0.9 | 0.3 |
|  | 35-49 | 202974 | 5.5 | 2.5 | 0.5 | 1.7 | 0.4 | 3.0 | 1.2 | 1.3 | 0.6 |
|  | 50-64 | 189493 | 8.9 | 2.4 | 0.5 | 1.3 | 0.5 | 6.4 | 2.8 | 2.5 | 1.2 |
|  | 65-74 | 90736 | 18.1 | 2.3 | 0.6 | 1.1 | 0.6 | 15.8 | 7.3 | 5.8 | 2.7 |
|  | 75-84 | 62205 | 32.9 | 3.3 | 0.8 | 1.8 | 0.7 | 29.6 | 14.8 | 10.4 | 4.4 |
|  | 85+ | 22570 | 71.9 | 4.9 | 1.8 | 2.2 | 0.9 | 67.0 | 34.2 | 23.3 | 9.6 |
|  | 18-64 | 672099 | 6.2 | 2.8 | 0.4 | 1.8 | 0.5 | 3.4 | 1.3 | 1.4 | 0.6 |
|  | 65+ | 175502 | 30.3 | 3.0 | 0.8 | 1.5 | 0.7 | 27.3 | 13.4 | 9.7 | 4.2 |
